# OCD symptom severity and comorbid psychiatric diagnoses in a Swedish genetic epidemiological obsessive-compulsive disorder cohort

**DOI:** 10.1101/2021.06.28.21259652

**Authors:** Behrang Mahjani, Christina Gustavsson Mahjani, Abraham Reichenberg, Sven Sandin, Christina M. Hultman, Joseph D. Buxbaum, Dorothy E. Grice

## Abstract

**Background:** We have established an epidemiological obsessive-compulsive disorder (OCD) cohort in Sweden. Individuals contributed DNA for genotyping and sequencing and also completed a Swedish translation of the Obsessive-Compulsive Inventory-Revised (OCI-R), a self-report questionnaire for assessing the severity and type of symptoms of OCD. This study made use of the OCI-R data to examine the severity and symptom dimensions of OCD as well as comorbidity with other psychiatric disorders.

**Methods:** OCI-R data for 1,134 individuals were available for this study, 1,010 diagnosed with OCD, and 124 diagnosed with chronic tic disorders without OCD used as a comparison group. We first evaluated the psychometric properties of the Swedish translation of the OCI-R. Then, we linked data from the Swedish national registries to access and analyze psychiatric comorbidities of OCD.

**Results:** The Swedish translation of OCI-R demonstrated internal consistency (Cronbach’s α = 0.9) and clear agreement with the OCI-R six-factor model. The mean total OCI-R score for females was significantly higher than for males. The most comorbid psychiatric condition to OCD were anxiety disorders (13.6%) and major depression (12%). We observed that individuals with OCD frequently had additional comorbid psychiatric disorders and that the severity of OCD was significantly higher in individuals with at least one additional psychiatric comorbidity compared to individuals with no psychiatric comorbidity.

**Conclusion:** We showed that the Swedish translation of the OCI-R has appropriate psychometric properties. Using an epidemiological framework, we were able to assess the severity and symptom dimensions of OCD and comorbidity with other psychiatric disorders.

## 1. Introduction

Obsessive-compulsive disorder (OCD) is a common psychiatric disorder characterized by intrusive and unwanted thoughts, images, or urges (obsessions) and repetitive behaviors (compulsions) that function to reduce the distress caused by obsessions. The prevalence of OCD is estimated at 0.75-2.5% of the general population (Fontenelle, Mendlowicz, & Versiani, 2006; Karno, Golding, Sorenson, & Burnam, 1988; Behrang Mahjani et al., 2020; Osland, Arnold, & Pringsheim, 2018; Ruscio, Stein, Chiu, & Kessler, 2010; Torres, Albina et al., 2006; Weissman, Bland, & Canino, 1994).

The Obsessive-Compulsive Inventory-Revised (OCI-R) is a self-report questionnaire that is designed to assess the severity and type of symptoms that are typically present in OCD (Foa et al., 2002). By answering 18 questions, individuals rate the degree to which they are bothered or distressed during the past month by specific, common OCD symptoms, using a 5-point scale from not at all (0 points) to extremely (4 points). The OCI-R measures six dimensions of OCD symptoms: ordering, obsessing, checking, cleaning, hoarding, and neutralizing. The OCI-R correlates with the widely-accepted OCD severity rating assessment scale, the Yale-Brown Obsessive-Compulsive Scale (Y-BOCS) (Abramowitz & Deacon, 2006; Goodman et al., 1989; Storch et al., 2010). However, OCI-R covers a subset of the scales included in Y-BOCS and does not directly measure time occupied, interference, resistance, or control related to OCD (the remaining Y-BOCS subscales).

The OCI-R is a well-established instrument with excellent psychometric properties (Foa et al., 2002). It has high test-retest reliability, convergent validity, good internal consistency, and solid factor structure according to both clinical and non-clinical studies (Gönner, Leonhart, & Ecker, 2008; Grabill et al., 2008; Hajcak, Huppert, Simons, & Foa, 2004; Huppert et al., 2007). The translated version of OCI-R into Norwegian (Solem, Hjemdal, Vogel, & Stiles, 2010), Spanish (Fullana et al., 2005), German (Gönner et al., 2008), Icelandic (Smárt,Ólason, Eypórsdóttir, & Frölunde, 2007), and Chinese (Hon, Siu, Cheng, Wong, & Foa, 2019) had shown satisfactory psychometric properties. A Swedish translation of the OCI-R has been available since 2009. Despite the extensive use in Swedish clinical settings, no evaluation of the psychometric properties has been made previously. The goal of this study was to evaluate the psychometric properties of the Swedish OCI-R and shed light on the frequency, severity, and symptom dimensions of OCD comorbid with other psychiatric conditions.

## 2. Method

### 2.1 Translation

The OCI-R was translated into Swedish by Anna Grönberg and Sandra Bates in 2009, based on the original OCI-R questionnaire Foa *et al*. (Foa et al., 2002), with permission from the original authors and without copyright.

### 2.2 Study population

In this study, we used data collected from study participants in EGOS (Epidemiology and Genetics of Obsessive-Compulsive Disorder and Chronic Tic Disorders in Sweden), one of the largest ongoing population-based cohort studies in Sweden, with the overall aim to identify genetic and possible environmental risk factors for OCD and chronic tic disorders (B. Mahjani et al., 2020). The EGOS study has been led by Professor Dorothy Grice and is the result of a collaboration between Icahn School of Medicine at Mount Sinai, New York, USA, and Karolinska Institutet, Stockholm, Sweden. Ethical approval was obtained from the Institutional Review Board at the Icahn School of Medicine at Mount Sinai, New York, USA, and the Regional Ethical Review Board in Stockholm, Sweden.

EGOS is built upon clinical diagnoses of OCD and CTD in specialized psychiatric care by the treating physician. In the EGOS study, all individuals living in Sweden with a clinical diagnosis of OCD or CTD by a psychiatrist in the Swedish National Patient Register (NPR) were eligible for inclusion in the source population. In NPR, the diagnoses were recorded using the International Classification of Disease (ICD) criteria. Individuals with at least two clinical diagnoses of OCD or CTD in the source population were contacted to join the study and provide either a blood sample or saliva. DNA was extracted upon arrival of the samples at the Biobank. All samples were genotyped using the Global Screening Array and analyzed for common genetic variation (B. Mahjani et al., 2021). All samples are currently going through whole-exome sequencing to detect rare structural variation and single nucleotide variation. The Swedish translation of OCI-R was provided as a web questionnaire to all participants (for more details, see the EGOS cohort profile (B. Mahjani et al., 2020)).

Our study population in this work consisted of 1,134 individuals who had completed the Swedish translation of the OCI-R questionnaire by June 2019. The date of the first registered diagnosis in the NPR was used as the diagnosis date.

### 2.3 Statistical analysis

We used the *psych* package developed by William Revelle in R (Revelle, 2019). We performed a confirmatory factor analysis to confirm the six-factor structure of OCI-R and address the construct validity. We used the criteria recommended by Hu and Bentler (Hu L.-T. & Bentler P. M., 1999): 1) *χ*^*2*^*-*statistic and the p-value: a non-significant result suggests that the model fits. The statistic becomes most likely significant for large samples; 2) Comparative fit index (CFI): should be ≥ 0.95; 3) Root mean squared error of approximation (RMSEA) including 90% CI: RMSEA should be ≤ 0.05, upper confidence interval (CI) bound ≤ 0.10; 4) Standardized root mean square residual (SRMR): should be ≤ 0.08 (Mair, 2018).

For internal consistency reliability, we estimated Cronbach’s α coefficient. Cronbach’s α reflects how closely related a set of test items are as a group.

OCI-R score is sensitive to treatment effects, and pre-to post changes in OCI-R score could reflect an improvement in OCD (Abramowitz, Tolin, & Diefenbach, 2005). Given that our source population was sampled from those in specialized psychiatric care, we anticipate that most individuals received some treatment. We analyzed the association between the latest diagnosis date in the NPR and the date the individual completed the OCI-R questionnaire (we refer to it as *time difference*). We adjusted the total OCI-R score by using the time difference as a covariate in linear regression. We also adjusted Cronbach’s α coefficient using multi-facet G-theory and applying the time difference as a facet.

## 3. Results

### 3.1 Demographic Data and frequency of comorbid psychiatric conditions

OCI-R data for 1,134 individuals were available for this study. Among 1,134 individuals, 1,010 had a diagnosis of OCD with an average diagnosis age of 22.1 (SD=7.0). OCD was more common among females than males (65% vs. 35%). We explored the frequency of comorbid psychiatric disorders with OCD. Anxiety disorders (13.6%) and major depressive disorder (12%) were the most common comorbid diagnoses (Table 1). 124 individuals had CTD without OCD (42 females and 82 males).

**Table 1.**
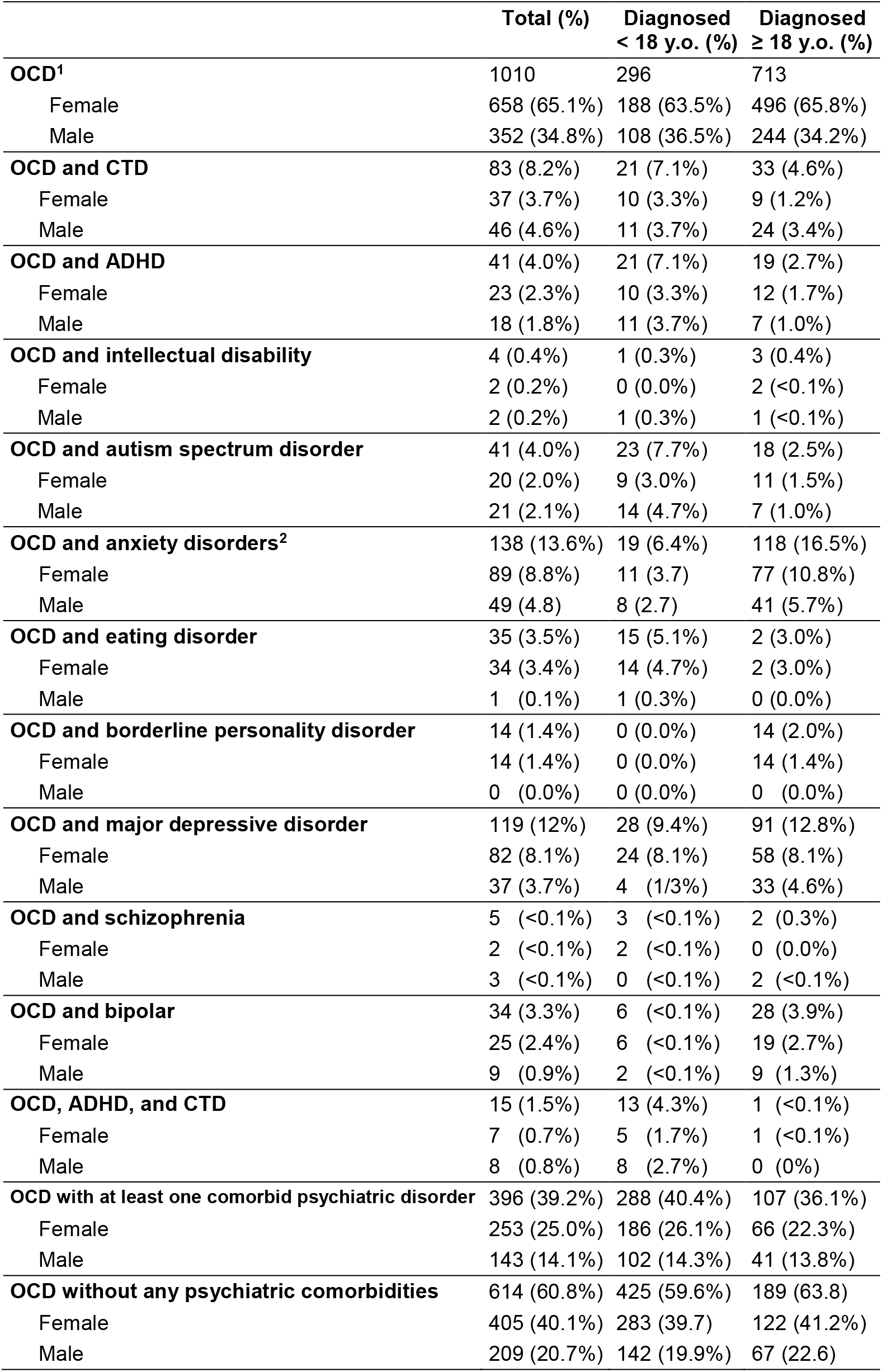
Characterization of the comorbid conditions with OCD

|  | **Total (%)** | **Diagnosed**<br>**< 18 y.o. (%)** | **Diagnosed**<br>**≥ 18 y.o. (%)** |
| --- | --- | --- | --- |
| **OCD<sup>1</sup>** | 1010 | 296 | 713 |
| Female | 658 (65.1%) | 188 (63.5%) | 496 (65.8%) |
| Male | 352 (34.8%) | 108 (36.5%) | 244 (34.2%) |
| **OCD and CTD** | 83 (8.2%) | 21 (7.1%) | 33 (4.6%) |
| Female | 37 (3.7%) | 10 (3.3%) | 9 (1.2%) |
| Male | 46 (4.6%) | 11 (3.7%) | 24 (3.4%) |
| **OCD and ADHD** | 41 (4.0%) | 21 (7.1%) | 19 (2.7%) |
| Female | 23 (2.3%) | 10 (3.3%) | 12 (1.7%) |
| Male | 18 (1.8%) | 11 (3.7%) | 7 (1.0%) |
| **OCD and intellectual disability** | 4 (0.4%) | 1 (0.3%) | 3 (0.4%) |
| Female | 2 (0.2%) | 0 (0.0%) | 2 (<0.1%) |
| Male | 2 (0.2%) | 1 (0.3%) | 1 (<0.1%) |
| **OCD and autism spectrum disorder** | 41 (4.0%) | 23 (7.7%) | 18 (2.5%) |
| Female | 20 (2.0%) | 9 (3.0%) | 11 (1.5%) |
| Male | 21 (2.1%) | 14 (4.7%) | 7 (1.0%) |
| **OCD and anxiety disorders<sup>2</sup>** | 138 (13.6%) | 19 (6.4%) | 118 (16.5%) |
| Female | 89 (8.8%) | 11 (3.7) | 77 (10.8%) |
| Male | 49 (4.8) | 8 (2.7) | 41 (5.7%) |
| **OCD and eating disorder** | 35 (3.5%) | 15 (5.1%) | 2 (3.0%) |
| Female | 34 (3.4%) | 14 (4.7%) | 2 (3.0%) |
| Male | 1 (0.1%) | 1 (0.3%) | 0 (0.0%) |
| **OCD and borderline personality disorder** | 14 (1.4%) | 0 (0.0%) | 14 (2.0%) |
| Female | 14 (1.4%) | 0 (0.0%) | 14 (1.4%) |
| Male | 0 (0.0%) | 0 (0.0%) | 0 (0.0%) |
| **OCD and major depressive disorder** | 119 (12%) | 28 (9.4%) | 91 (12.8%) |
| Female | 82 (8.1%) | 24 (8.1%) | 58 (8.1%) |
| Male | 37 (3.7%) | 4 (1/3%) | 33 (4.6%) |
| **OCD and schizophrenia** | 5 (<0.1%) | 3 (<0.1%) | 2 (0.3%) |
| Female | 2 (<0.1%) | 2 (<0.1%) | 0 (0.0%) |
| Male | 3 (<0.1%) | 0 (<0.1%) | 2 (<0.1%) |
| **OCD and bipolar** | 34 (3.3%) | 6 (<0.1%) | 28 (3.9%) |
| Female | 25 (2.4%) | 6 (<0.1%) | 19 (2.7%) |
| Male | 9 (0.9%) | 2 (<0.1%) | 9 (1.3%) |
| **OCD, ADHD, and CTD** | 15 (1.5%) | 13 (4.3%) | 1 (<0.1%) |
| Female | 7 (0.7%) | 5 (1.7%) | 1 (<0.1%) |
| Male | 8 (0.8%) | 8 (2.7%) | 0 (0%) |
| **OCD with at least one comorbid psychiatric disorder** | 396 (39.2%) | 288 (40.4%) | 107 (36.1%) |
| Female | 253 (25.0%) | 186 (26.1%) | 66 (22.3%) |
| Male | 143 (14.1%) | 102 (14.3%) | 41 (13.8%) |
| **OCD without any psychiatric comorbidities** | 614 (60.8%) | 425 (59.6%) | 189 (63.8) |
| Female | 405 (40.1%) | 283 (39.7) | 122 (41.2%) |
| Male | 209 (20.7%) | 142 (19.9%) | 67 (22.6) |

We compared the characteristics of individuals with OCD who were diagnosed before the age of 18 (early diagnosis age) and after the age of 18 (late diagnosis age). In both age categories, OCD was more common in females than males. Among individuals with late diagnosis age, anxiety disorders, major depressive disorder, borderline personality disorder, and bipolar disorder were the most common psychiatric comorbidities. In contrast, among individuals with early diagnosis age, Tourette disorder and other CTD, ADHD, autism spectrum disorder, and eating disorder were most common.

39.2% of individuals with OCD had at least one additional psychiatric comorbidity, with about a similar rate for individuals diagnosed before or after the age of 18.

### 3.2 Psychometric properties and construct validity

#### 3.2.1 Confirmatory factor analysis

We performed a confirmatory factor analysis with six factors: Washing, Obsessing, Hoarding, Ordering, Checking, and Neutralizing. We estimated χ2 (120) = 401.056 (p = 0.000), RMSEA = 0.048 (90% CI, 0.043, 0.053), TLI = 0.996, SRMR = 0.043 and CFI = 0.997. According to these values, the six-factor structure of the original version of the OCI-R was supported, based on the criteria recommended by Hu and Bentler (Hu L.-T. & Bentler P. M., 1999).

The model of the confirmatory factor analysis is illustrated in Figure 1. The standardized factor weights are shown on the arrows. The correlations between the factors were between 0.39 and 0.52, indicating that the subscales were related to each other but not redundant.

**Figure 1.**
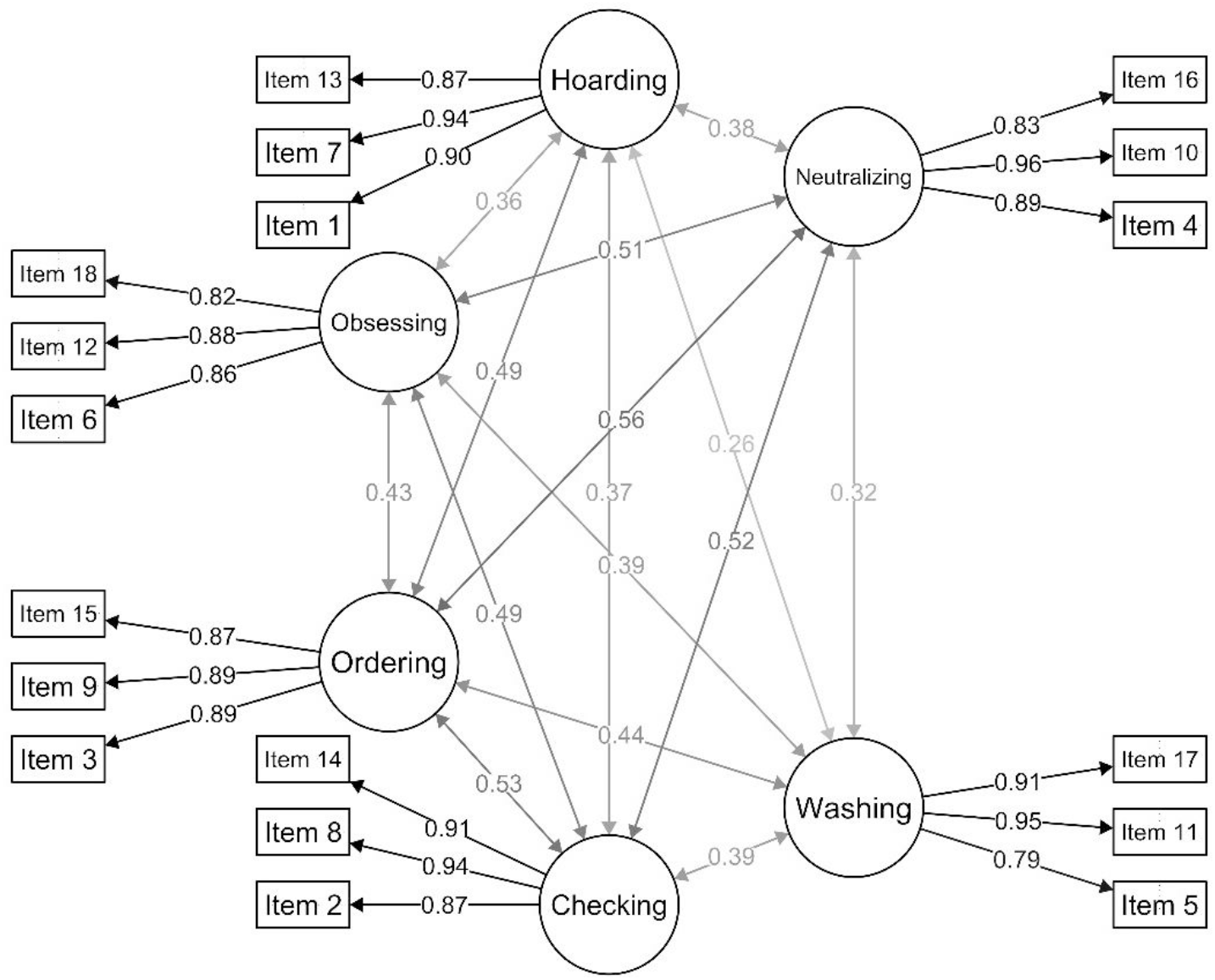
Model of the confirmatory factor analysis for the Swedish translation of the Obsessive-Compulsive Inventory-Revised. All estimates are standardized. Rectangles represent latent variables (questions in OCI-R). Circles represent factors. Correlations between factors are shown by double-headed arrows. Values next to arrows between the latent variables (questions) and the factors are standardized regression weights. The closer they are to one, the larger they are to account for the variation in that factor.

#### 3.2.2 Internal consistency

The Cronbach’s α coefficient for the total score was 0.9 [95% CI, 0.89-0.91]. The results for the subscales were: Washing, α = 0.88; Obsessing, α = 0.85; Hoarding, α = 0.87; Ordering, α = 0.88; Checking, α = 0.89; and Neutralizing, α = 0.87 (Table 3). The α values were close to the reported α values for the original OCI-R (Foa et al., 2002). We also used G-theory to calculate the generalizability coefficient using variance components, a reliability coefficient equivalent to Cronbach’s α. The generalizability coefficient decreased slightly after adjusting for the time differences between the latest diagnosis date and the date that individuals completed the OCI-R (using time differences as facet) (Table 2).

**Table 2.** Internal consistency for the Swedish translation of the Obsessive-Compulsive Inventory-Revised.

|  | OCD | CTD without OCD | OCD |  |
| --- | --- | --- | --- | --- |
| | Cronbach's $\alpha$ coefficient<br>(95% CI) | Cronbach's $\alpha$ coefficient<br>(95% CI) | Generalizability<br>before<br>adjustment | Generalizability<br>after<br>adjustment |
| **Total score** | 0.90 (0.89-0.91) | 0.85 (0.81-0.89) | 0.900 | 0.897 |
| **Obsessing** | 0.85 (0.84-0.87) | 0.85 (0.80-0.89) | 0.851 | 0.848 |
| **Washing** | 0.88 (0.87-0.89) | 0.80 (0.73-0.86) | 0.882 | 0.876 |
| **Hoarding** | 0.87 (0.86-0.88) | 0.70 (0.61-0.80) | 0.870 | 0.870 |
| **Ordering** | 0.88 (0.87-0.89) | 0.82 (0.76-0.87) | 0.881 | 0.879 |
| **Checking** | 0.89 (0.87-0.90) | 0.83 (0.78-0.88) | 0.887 | 0.887 |
| **Neutralizing** | 0.87 (0.86-0.89) | 0.70 (0.60-0.79) | 0.874 | 0.872 |

**Table 3.** OCI-R score for individuals with OCD and other comorbid conditions

|  | Total OCI-R score (SD) |
| --- | --- |
| **OCD** | 22.5 (13.6) |
| Female | 23.3 (13.9) |
| Male | 20.9 (12.8) |
| **OCD and CTD** | 20.0 (13.2) |
| Female | 21.4 (15.0) |
| Male | 18.9 (11.6) |
| **OCD and ADHD** | 25.5 (15.7) |
| Female | 31.1 (16.1) |
| Male | 18.3 (12.2) |
| **OCD and intellectual disability** | 31.8 (21.3) |
| Female | 49.5 (4.9) |
| Male | 14.0 (8.5) |
| **OCD and autism spectrum disorder** | 22.0 (12.6) |
| Female | 21.4 (12.5) |
| Male | 22.6 (13.1) |
| **OCD and anxiety disorders<sup>1</sup>** | 23.9 (12.9) |
| Female | 24.2 (13.9) |
| Male | 23.3 (11.1) |
| **OCD and eating disorder** | 24.3 (16.1) |
| Female | 24.3 (16.4) |
| Male | 26.0 (-) |
| **OCD and borderline personality disorder** | 29.2 (13.6) |
| Female | 29.2 (13.6) |
| Male | 0 |
| **OCD and major depressive disorder** | 25.3 (15.9) |
| Female | 26.9 (16.1) |
| Male | 21.8 (15.1) |
| **OCD and schizophrenia** | 14.6 (11.7) |
| Female | 17.7 (15.4) |
| Male | 10.0 (1.41) |
| **OCD and bipolar** | 29.5 (14.2) |
| Female | 31.1 (14.6) |
| Male | 25.1 (12.6) |
| **OCD, ADHD, and CTD** | 19.47 (17.6) |
| Female | 30.57 (19.7) |
| Male | 9.75 (7.5) |
| **CTD without OCD** | 14.3 (9.8) |
| Female | 17.7 (11.4) |
| Male | 12.6 (8.5) |
| **OCD with at least one comorbid psychiatric disorder** | 24.1 (14.1) |
| Female | 25.4 (14.8) |
| Male | 21.9 (12.5) |
| **OCD without any psychiatric comorbidities** | 21.4 (13.1) |
| Female | 22.0 (13.2) |
| Male | 20.3 (12.9) |
| **OCD diagnosed < 18 y.o.** | 20.3 (12.7) |
| Female | 21.9 (13.6) |
| Male | 17.6 (10.7) |
| **OCD diagnosed ≥ 18 y.o.** | 23.4 (13.8) |
| Female | 23.9 (14.0) |
| Male | 22.4 (13.3) |

<sup>1</sup>Phobic anxiety disorders (F40), and other anxiety disorders (F41).

We calculated Cronbach’s α coefficient for individuals with CTD without OCD (Table 2). For this group, Cronbach’s α coefficients were closer to the control populations reported in the other studies (Solem et al., 2010).

### 3.3 Population characteristics of obsessive-compulsive symptoms

The mean total OCI-R score for individuals with OCD was 22.5 (SD = 13.6) and after adjusting for the time difference was 29.0 (SD = 13.0)(Table 3). In addition, females with OCD had significantly higher OCI-R scores compared to males (23.3 vs. 20.9, p-value < 0.01).

The mean total OCI-R score for individuals with CTD and without OCD was 14.3 (SD=9.8), and it was slightly higher than the control populations reported in the other studies (8.3 (SD=9.2) in (Sica et al., 2009), and 10.4 (SD=9.2) in (Solem et al., 2010)). OCI-R score of individuals with OCD was significantly higher than individuals with CTD without OCD (22.1 vs. 14.3, p-value < 0.01).

Individuals with OCD and at least one comorbid psychiatric disorder had significantly higher OCI-R scores than individuals with OCD without any psychiatric comorbidity (24.1 Vs. 21.4, p-value < 0.01). In particular, the mean total OCI-R score for individuals with OCD and bipolar (OCI-R = 29.5, p-value < 0.01), OCD and borderline personality disorder (OCI-R = 29.2, p-value = 0.03), OCD and major depressive disorder (OCI-R = 25.3, p-value = 0.007), and OCD and anxiety disorders (OCI-R = 23.9, p-value = 0.02) were significantly higher than individuals with OCD without any psychiatric comorbidity (OCI-R = 21.4).

Individuals with late OCD diagnosis age (≥ 18 y.o.) had significantly higher OCI-R score than individuals with early OCD diagnosis age (< 18 y.o.) (OCI-R 23.4 vs. 20.3, p-value<0.001).

We compared the OCI-R subscales of individuals with different comorbid psychiatric conditions (Table 4 and Appendix, Table A1). Scores for Washing, Obsessing, Checking, and Neutralizing of individuals with OCD were significantly different from the scores of individuals with CTD without OCD (p-value < 0.01). For Ordering and Hoarding, the differences were not significant.

**Table 4.** OCI-R subscales

|  | **Washing**<br>**(SD)** | **Obsessing**<br>**(SD)** | **Hoarding**<br>**(SD)** | **Ordering**<br>**(SD)** | **Checking**<br>**(SD)** | **Neutralizing**<br>**(SD)** |
| --- | --- | --- | --- | --- | --- | --- |
| **OCD** | 4.01 (3.68) | 5.29 (3.36) | 2.08 (2.66) | 3.93 (3.35) | 4.34 (3.33) | 2.82 (3.44) |
| Female | 4.34 (3.80) | 5.34 (3.33) | 2.05 (2.69) | 4.18 (3.46) | 4.51 (3.37) | 2.88 (3.46) |
| Male | 3.39 (3.36) | 5.20 (3.43) | 2.13 (2.59) | 3.47 (3.08) | 4.04 (3.24) | 2.70 (3.41) |
| **CTD without OCD** | 1.55 (2.41) | 3.22 (2.95) | 2.27 (2.18) | 3.55 (3.05) | 2.47 (2.59) | 1.25 (2.13) |
| Female | 1.81 (2.34) | 3.71 (3.40) | 2.57 (2.54) | 4.55 (3.33) | 3.14 (2.93) | 1.88 (2.72) |
| Male | 1.41 (2.45) | 2.96 (2.68) | 2.12 (1.98) | 3.04 (2.78) | 2.12 (2.35) | 0.93 (1.68) |

## 4. Discussion

In this study, for the first time, we demonstrated that the Swedish translation of OCI-R has adequate psychometric properties. We found support for the six-factor structure of OCI-R: Washing, Obsessing, Hoarding, Ordering, Checking, and Neutralizing. To evaluate the internal consistency of the Swedish OCI-R, we estimated the Cronbach α value. Cronbach α value for the total score was 0.9, and smaller than 0.9 for all the subscales. We calculated the generalizability coefficient and adjusted for the time differences between the latest diagnosis date and the date that the individuals completed the OCI-R. We did not observe a substantial change in the Cronbach α value before and after adjusting for the time differences.

The OCI-R total score was slightly lower than the reported results in other studies (22.5 in this study vs. 25 in (Sica et al., 2009) and 30.1 in (Solem et al., 2010)). However, we showed that the time difference between the most recent diagnosis date and the date the individual completed the questionnaire, likely reflecting time in treatment, could explain the smaller estimates (22.5 before adjustment vs. 29.0 after adjustment).

A unique feature of the EGOS data is the availability of information about all comorbid psychiatric conditions. We observed that 13.6% of individuals with OCD had a diagnosis of anxiety disorders, and 12% major depressive disorder. Individuals with OCD and bipolar disorder had the highest OCI-R total score in comparison to other comorbidities. The severity of OCD was significantly higher in individuals with at least one psychiatric comorbidity than individuals with no psychiatric comorbidity. We conjecture that individuals with more severe symptoms carry a larger genetic load, which is the goal of our next study.

The present study had several strengths and some limitations: 1) we used data from the Swedish NPR, which created a genetically homogeneous sample and would be expected to minimize the risk of confounding due to population stratification; 2) all individuals had a clinical diagnosis of OCD by a specialist, which minimizes misdiagnosis; 3) a control group consisting of healthy individuals (individuals with no psychiatric diagnosis) was not available for this study. However, data were available for a group of individuals with CTD without OCD. For this group, Cronbach’s α coefficient was close to the control populations reported by other studies (Solem et al., 2010). Due to the genetic and phenotypic correlation of OCD and CTD, this group could not be treated as a control group, although the contrast between the results was informative; 4) another limitation of this study was the use of data from NPR. While the NPR is considered a robust and reliable source for research, utilizing ICD diagnostic criteria, the register is lacking information about the individuals who do not seek clinical services at all or are treated at primary care. Hence, our sample may over-represented more severe cases.

## Data Availability

Study data are maintained at the Department of Medical Epidemiology and Biostatistics, Karolinska Institutet,
Stockholm, Sweden. Extracted DNA samples are stored at Karolinska Biobank [https ://ki.se/en/resea rch/ki-bioba
nk]. Biological samples can be made available to approved researchers.
EGOS informed consents permit sharing of genetic data via the database of Genotypes and Phenotypes (dbGaP).

## Acknowledgments

This study was supported by a grant from the Beatrice and Samuel A. Seaver Foundation (DEG, JDB, BM); the Mindworks Charitable Lead Trust (DEG); the Stanley Center for Psychiatric Research (DEG); NIMH R01MH124679 (DEG).

## Appendix

**Table A1.** OCI-R subscales

|  | **Washing**<br>**(SD)** | **Obsessing**<br>**(SD)** | **Hoarding**<br>**(SD)** | **Ordering**<br>**(SD)** | **Checking**<br>**(SD)** | **Neutralizing**<br>**(SD)** |
| --- | --- | --- | --- | --- | --- | --- |
| **OCD** | 4.01 (3.68) | 5.29 (3.36) | 2.08 (2.66) | 3.93 (3.35) | 4.34 (3.33) | 2.82 (3.44) |
| Female | 4.34 (3.80) | 5.34 (3.33) | 2.05 (2.69) | 4.18 (3.46) | 4.51 (3.37) | 2.88 (3.46) |
| Male | 3.39 (3.36) | 5.20 (3.43) | 2.13 (2.59) | 3.47 (3.08) | 4.04 (3.24) | 2.70 (3.41) |
| **OCD and CTD** | 3.16 (3.00) | 4.61 (3.29) | 1.98 (2.21) | 3.69 (3.54) | 3.69 (3.31) | 2.86 (3.71) |
| Female | 3.41 (3.40) | 5.14 (3.58) | 2.19 (2.48) | 4.14 (4.02) | 3.89 (3.36) | 2.59 (3.56) |
| Male | 2.96 (2.66) | 4.20 (3.01) | 1.80 (1.98) | 3.33 (3.10) | 3.52 (3.29) | 3.07 (3.85) |
| **OCD and ADHD** | 4.39 (3.90) | 5.95 (3.86) | 2.56 (2.92) | 4.66 (3.66) | 4.93 (3.77) | 2.98 (3.61) |
| Female | 5.17 (4.30) | 7.00 (3.54) | 3.43 (3.00) | 5.70 (4.00) | 5.52 (3.84) | 4.26 (4.15) |
| Male | 3.39 (3.15) | 4.61 (3.93) | 1.44 (2.45) | 3.33 (2.74) | 4.17 (3.65) | 1.33 (1.81) |
| **OCD and intellectual disability** | 4.75 (4.57) | 6.75 (3.40) | 4.75 (1.71) | 6.25 (4.03) | 5.25 (5.74) | 4.00 (5.66) |
| Female | 7.50 (4.95) | 9.00 (1.41) | 6.00 (1.41) | 9.00 (4.24) | 10.0 (2.83) | 8.00 (5.66) |
| Male | 2.00 (2.83) | 4.50 (3.54) | 3.50 (0.71) | 3.50 (0.71) | 0.50 (0.71) | 0.00 (0.00) |
| **OCD and autism spectrum disorder** | 4.15 (3.38) | 5.73 (3.66) | 2.68 (2.87) | 3.63 (2.96) | 3.44 (3.00) | 2.37 (2.92) |
| Female | 4.60 (3.28) | 5.65 (3.33) | 2.15 (2.81) | 3.20 (3.24) | 3.20 (2.78) | 2.60 (2.93) |
| Male | 3.71 (3.49) | 5.81 (4.03) | 3.19 (2.89) | 4.05 (2.69) | 3.67 (3.25) | 2.14 (2.97) |
| **OCD and anxiety disorders<sup>1</sup>** | 4.36 (3.51) | 5.96 (3.42) | 2.20 (2.89) | 4.38 (3.34) | 3.53 (3.55) | 2.66 (3.31) |
| Female | 4.51 (3.63) | 5.74 (3.60) | 2.27 (3.19) | 4.73 (3.51) | 4.27 (3.52) | 2.73 (3.24) |
| Male | 4.10 (3.30) | 6.35 (3.05) | 2.06 (2.29) | 3.76 (2.95) | 4.47 (3.73) | 2.53 (3.45) |
| **OCD and eating disorder** | 4.97 (3.86) | 5.69 (3.75) | 2.34 (3.24) | 4.20 (3.63) | 4.57 (3.78) | 2.54 (3.12) |
| Female | 4.97 (3.92) | 5.50 (3.64) | 2.35 (3.28) | 4.24 (3.68) | 4.62 (3.83) | 2.59 (3.15) |
| Male | 5 (-) | 12 (-) | 2 (-) | 3 (-) | 3 (-) | 1 (-) |
| **OCD and borderline personality disorder** | 6.29 (3.27) | 5.79 (3.72) | 1.71 (2.43) | 5.79 (3.38) | 5.00 (3.86) | 4.64 (3.23) |
| Female | 6.29 (3.27) | 5.79 (3.72) | 1.71 (2.43) | 5.79 (3.38) | 5.00 (3.86) | 4.64 (3.23) |
| Male | - | - | - | - | - | - |
| **OCD and major depressive disorder** | 4.17 (3.83) | 5.94 (3.65) | 2.66 (3.16) | 4.30 (3.75) | 4.61 (3.52) | 3.62 (3.92) |
| Female | 4.62 (3.97) | 5.95 (3.68) | 2.76 (3.24) | 4.74 (3.82) | 4.89 (3.60) | 3.90 (3.88) |
| Male | 3.16 (3.34) | 5.92 (3.63) | 2.43 (3.01) | 3.32 (3.44) | 3.97 (3.30) | 3.00 (3.99) |
| **OCD and schizophrenia** | 1.80 (1.64) | 3.60 (2.30) | 1.60 (1.67) | 2.20 (2.17) | 3.00 (2.65) | 2.40 (3.91) |
| Female | 1.67 (2.08) | 3.33 (3.06) | 2.00 (2.00) | 3.00 (2.65) | 3.67 (3.51) | 4.00 (4.58) |
| Male | 2.00 (1.41) | 4.00 (1.41) | 1.00 (1.41) | 1.00 (-) | 2.00 (-) | 0 (-) |
| **OCD and Bipolar** | 4.53 (3.74) | 6.32 (3.57) | 2.71 (3.56) | 6.74 (3.73) | 5.56 (3.63) | 3.65 (3.82) |
| Female | 5.36 (3.81) | 6.44 (3.47) | 2.28 (3.32) | 6.96 (3.92) | 6.24 (3.48) | 3.80 (4.13) |
| Male | 2.22 (2.44) | 6.00 (4.06) | 3.89 (4.14) | 6.11 (3.26) | 3.67 (3.54) | 3.22 (2.95) |
| **OCD with at least one comorbid psychiatric disorder** | 4.14 (3.61) | 5.77 (3.51) | 2.36 (2.86) | 4.45 (3.51) | 4.38 (3.45) | 3.04 (3.59) |
| Female | 4.57 (3.78) | 5.81 (3.58) | 2.37 (2.96) | 4.86 (3.65) | 4.58 (3.50) | 3.21 (3.64) |
| Male | 3.35 (3.15) | 5.69 (3.40) | 2.35 (2.69) | 3.72 (3.14) | 4.03 (3.33) | 2.73 (3.50) |
| **OCD without any psychiatric comorbidities** | 3.93 (3.73) | 4.98 (3.23) | 1.89 (2.51) | 3.60 (3.19) | 4.32 (3.25) | 2.68 (3.34) |
| Female | 4.20 (3.82) | 5.04 (3.13) | 1.85 (2.50) | 3.76 (3.26) | 4.46 (3.28) | 2.68 (3.34) |
| Male | 3.42 (3.50) | 4.86 (3.42) | 1.99 (2.52) | 3.29 (3.04) | 4.05 (3.19) | 2.67 (3.35) |
| **OCD diagnosed < 18 y.o.** | 4.02 (3.54) | 4.95 (3.41) | 1.95 (2.42) | 3.36 (2.98) | 3.70 (3.11) | 2.34 (3.11) |
|  | 4.32 (3.66) | 5.23 (3.41) | 1.99 (2.47) | 3.70 (3.18) | 4.10 (3.28) | 2.55 (3.31) |
|  | 3.50 (3.27) | 4.45 (3.39) | 1.87 (2.32) | 2.77 (2.50) | 2.99 (2.65) | 1.99 (2.71) |
| **OCD diagnosed ≥ 18 y.o.** | 4.01 (3.74) | 5.43 (3.33) | 2.12 (2.75) | 4.16 (3.46) | 4.61 (3.38) | 3.01 (3.56) |
| Female | 4.36 (3.87) | 5.38 (3.30) | 2.06 (2.77) | 4.36 (3.54) | 4.67 (3.39) | 3.01 (3.52) |
| Male | 3.34 (3.40) | 5.52 (3.40) | 2.25 (2.70) | 3.77 (3.26) | 4.51 (3.37) | 3.01 (3.64) |
| **CTD without OCD** | 1.55 (2.41) | 3.22 (2.95) | 2.27 (2.18) | 3.55 (3.05) | 2.47 (2.59) | 1.25 (2.13) |
| Female | 1.81 (2.34) | 3.71 (3.40) | 2.57 (2.54) | 4.55 (3.33) | 3.14 (2.93) | 1.88 (2.72) |
| Male | 1.41 (2.45) | 2.96 (2.68) | 2.12 (1.98) | 3.04 (2.78) | 2.12 (2.35) | 0.93 (1.68) |

<sup>1</sup>Phobic anxiety disorders (F40), and other anxiety disorders (F41).

## References

Abramowitz, J. S., & Deacon, B. J. (2006). Psychometric properties and construct validity of the Obsessive-Compulsive Inventory-Revised: Replication and extension with a clinical sample. Journal of Anxiety Disorders, 20(8), 1016–1035. https://doi.org/10.1016/j.janxdis.2006.03.001

Abramowitz, J. S., Tolin, D. F., & Diefenbach, G. J. (2005). Measuring change in OCD: Sensitivity of the obsessive-compulsive inventory-revised. Journal of Psychopathology and Behavioral Assessment, 27(4), 317–324. https://doi.org/10.1007/s10862-005-2411-y

Foa, E. B., Huppert, J. D., Leiberg, S., Langner, R., Kichic, R., Hajcak, G., & Salkovskis, P. M. (2002). The obsessive-compulsive inventory: Development and validation of a short version. Psychological Assessment, 14(4), 485–496. https://doi.org/10.1037/1040-3590.14.4.485

Fontenelle, L. F., Mendlowicz, M. V., & Versiani, M. (2006). The descriptive epidemiology of obsessive-compulsive disorder. Progress in Neuro-Psychopharmacology and Biological Psychiatry, 30(3). https://doi.org/10.1016/j.pnpbp.2005.11.001

Fullana, M. A., Tortella-Feliu, M., Caseras, X., Andión, Ó., Torrubia, R., & Mataix-Cols, D. (2005). Psychometric properties of the Spanish version of the Obsessive-Compulsive Inventory - Revised in a non-clinical sample. Journal of Anxiety Disorders, 19(8), 893–903. https://doi.org/10.1016/j.janxdis.2004.10.004

Gönner, S., Leonhart, R., & Ecker, W. (2008). The Obsessive-Compulsive Inventory-Revised (OCI-R): Validation of the German version in a sample of patients with OCD, anxiety disorders, and depressive disorders. Journal of Anxiety Disorders, 22(4), 734–749. https://doi.org/10.1016/j.janxdis.2007.07.007

Goodman, W. K., Price, L. H., Rasmussen, S. A., Mazure, C., Fleischmann, R. L., Hill, C. L., … Charney, D. S. (1989). The Yale-Brown Obsessive Compulsive Scale: I. Development, Use, and Reliability. Archives of General Psychiatry, 46(11), 1006–1011. https://doi.org/10.1001/archpsyc.1989.01810110048007

Grabill, K., Merlo, L., Duke, D., Harford, K. L., Keeley, M. L., Geffken, G. R., & Storch, E. A. (2008). Assessment of obsessive-compulsive disorder: A review. Journal of Anxiety Disorders, 22(1), 1–17. https://doi.org/10.1016/j.janxdis.2007.01.012

Hajcak, G., Huppert, J. D., Simons, R. F., & Foa, E. B. (2004). Psychometric properties of the OCI-R in a college sample. Behaviour Research and Therapy, 42(1), 115–123. https://doi.org/10.1016/j.brat.2003.08.002

Hon, S. K., Siu, B. W., Cheng, C., Wong, W. C., & Foa, E. B. (2019). Validation of the Chinese Version of Obsessive-Compulsive Inventory-Revised. East Asian Archives of Psychiatry, 29(4), 103–111. https://doi.org/10.12809/eaap1832

Hu L.-T., & Bentler P. M. (1999). Cutoff criteria for fit indexes in covariance structure analysis: conventional criteria versus new alternatives.. Structural Equation Modeling, 6(July 2012), 1–55.

Huppert, J. D., Walther, M. R., Hajcak, G., Yadin, E., Foa, E. B., Simpson, H. B., & Liebowitz, M. R. (2007). The OCI-R: Validation of the subscales in a clinical sample. Journal of Anxiety Disorders, 21(3), 394–406. https://doi.org/10.1016/j.janxdis.2006.05.006

Karno, M., Golding, J. M., Sorenson, S. B., & Burnam, M. A. (1988). The Epidemiology of Obsessive-Compulsive Disorder in Five US Communities. Archives of General Psychiatry, 45(12), 1094–1099. https://doi.org/10.1001/archpsyc.1988.01800360042006

Mahjani, B., Dellenvall, K., Grahnat, A.-C. S., Karlsson, G., Tuuliainen, A., Reichert, J., … Grice, D. E. (2020). Cohort profile: Epidemiology and Genetics of Obsessive–compulsive disorder and chronic tic disorders in Sweden (EGOS). Social Psychiatry and Psychiatric Epidemiology. https://doi.org/10.1007/s00127-019-01822-7

Mahjani, B., Klei, L., Mattheisen, M., Halvorsen, M. W., Reichenberg, A., Roeder, K., … Grice, D. E. (2021). The genetic architecture of obsessive-compulsive disorder: Alleles across the frequency spectrum contribute liability to OCD. MedRxiv. https://doi.org/10.1101/2021.01.26.21250409

Mahjani, Behrang, Klei, L., Hultman, C. M., Larsson, H., Devlin, B., Buxbaum, J. D., … Grice, D. E. (2020). Maternal Effects as Causes of Risk for Obsessive-Compulsive Disorder. In Biological Psychiatry (Vol. 87, pp. 1045–1051). https://doi.org/10.1016/j.biopsych.2020.01.006

Mair, P. (2018). Modern Psychometrics with R. Springer.

Osland, S., Arnold, P. D., & Pringsheim, T. (2018). The prevalence of diagnosed obsessive compulsive disorder and associated comorbidities: A population-based Canadian study. Psychiatry Research, 268, 137–142. https://doi.org/10.1016/j.psychres.2018.07.018

Revelle, W. (2019). psych: Procedures for Psychological, Psychometric, and Personality Research, 1–358. Retrieved from https://cran.r-project.org/package=psych

Ruscio, A. M., Stein, D. J., Chiu, W. T., & Kessler, R. C. (2010). The epidemiology of obsessive-compulsive disorder in the National Comorbidity Survey Replication. Molecular Psychiatry, 15(1), 53–63. https://doi.org/10.1038/mp.2008.94

Sica, C., Ghisi, M., Altoè, G., Chiri, L. R., Franceschini, S., Coradeschi, D., & Melli, G. (2009). The Italian version of the Obsessive Compulsive Inventory: Its psychometric properties on community and clinical samples. Journal of Anxiety Disorders, 23(2), 204–211. https://doi.org/10.1016/j.janxdis.2008.07.001

Smárt, J., Ólason, D. T., Eypórsdóttir, Á., & Frölunde, M. B. (2007). Psychometric properties of the obsessive compulsive inventory-revised among Icelandic college students. Scandinavian Journal of Psychology, 48(2), 127–133. https://doi.org/10.1111/j.1467-9450.2007.00574.x

Solem, S., Hjemdal, O., Vogel, P. A., & Stiles, T. C. (2010). A Norwegian version of the Obsessive-Compulsive Inventory-Revised: Psychometric properties. Scandinavian Journal of Psychology, 51(6), 509–516. https://doi.org/10.1111/j.1467-9450.2009.00798.x

Storch, E. A., Rasmussen, S. A., Price, L. H., Larson, M. J., Murphy, T. K., & Goodman, W. K. (2010). Development and psychometric evaluation of the yale-brown obsessive-compulsive scale-second edition. Psychological Assessment, 22(2), 223–232. https://doi.org/10.1037/a0018492

Torres Albina, R., Prince, M. J., Bebbington, P., Bhugra, D., Brugha, T. S., Farrell, M., … Singleton, N. (2006). Obsessive-Compulsive Disorder: Prevalence, Comorbidity, Impact, and Help-Seeking in the British National Psychiatric Morbidity Survey of 2000. American Journal of Psychiatry, 15(163), 1978–1985. https://doi.org/10.1080/0954026021000045921

Weissman, M., Bland, R., & Canino, G. (1994). The cross national epidemiology of obsessive compulsive disorder. J Clin Psychiatry, (55), 5–10.

